# Physical Injuries and Burns among Refugees in Lebanon: Implications for Programs and Policies

**DOI:** 10.1101/2021.09.27.21264058

**Authors:** Samar Al-Hajj, Moustafa Moustafa, Majed El Hechi, Mohamad A. Chahrour, Ali A. Nasrallah, Haytham Kaafarani

**Affiliations:** Faculty of Health Sciences, American University of Beirut, Beirut, Lebanon; University of Virginia, Richmond, USA; Department of Surgery, American University of Beirut Medical Center, Beirut, Lebanon; Division of Trauma, Emergency Surgery and Surgical Critical Care, Massachusetts General Hospital, Harvard Medical School, Boston, Massachusetts, USA

## Abstract

**Background:** Refugees are prone to injury due to often austere living conditions, social and economic disadvantages, and limited access to health care services in host countries. This study systematically quantified the prevalence of physical injuries and burns among the refugee community in Western Lebanon and examined injury characteristics, risk factors and outcomes.

**Method:** We conducted a cluster-based population survey across 21 camps in the Bekaa region of Lebanon from February to April 2019. A modified version of the ‘Surgeons Overseas Assessment of Surgical Need (SOSAS)’ tool v 3.0 was administered to the head of the refugee household and documented all injuries sustained by family members over the last 12 months. Descriptive and univariate regression analyses were performed to understand the association between variables.

**Results:** 750 heads of household were surveyed. 112 (14.9%) household sustained injuries in the past 12 months, 39 of which (34.9%) reported disabling injuries that affected their work and daily living. Most injuries occurred inside the tent (29.9%). A burn was sustained by at least one household member in 136 (18.1%) households. The majority (63.7%) of burns affected children under 5 years and were mainly due to boiling liquid (50%). Significantly more burns were reported in households where caregivers have the inability to lockout children while cooking (25.6% vs 14.9%, p-value=0.001). Similarly, households with unemployed head significantly had more reported burns (19.7% vs 13.3%, p-value=0.05). Nearly 16.1% of injured refugees were unable to seek health care due to lack of health insurance coverage and financial liability.

**Conclusion:** Refugees suffer injuries and burns with substantial human and economic repercussions on individuals, their families and the host healthcare system. Resources should be allocated to designing safe camps and implementing educational and awareness programs with special focus on heating and cooking methods.

## Introduction

Injury is the leading cause of death and disability in people under 44 [1, 2]. While high income countries (HIC) demonstrated a steady decrease in injuries through time, low- and middle-income countries (LMICs) showed an increasing trend [3]. This is due to multiple reasons, including the absence of safety regulations and injury prevention strategies [4] in many LMICs.

The Eastern Mediterranean Region (EMR) has the highest rate of injuries related deaths and disability-adjusted life years (DALYs) among LMICs, particularly injury related to road traffic crashes and violence [5-7]. According to the Global Burden of Disease (GBD 2019), injury related deaths in the EMR were estimated at 56.2 per 100,000, ranking 4^th^ compared to global rates [8]. Wars and regional conflicts have exacerbated injury prevalence in many EMR countries and rendered the provision of health care services limited, if not, scarce, particularly in war-affected regions [9]. The recent Syrian conflict has resulted in what has been classified as the worst humanitarian crisis in history [10], with millions of individuals displaced internally or seeking refuge in bordering countries [11].

Lebanon has taken in the highest number of Syrian refugees relative to its population, estimated at nearly 30% of its current population [12]. The refugee crisis has strained the already fragile Lebanese healthcare system. Several studies have demonstrated a high prevalence of communicable disease [10, 13-15] and injuries of varying etiologies among the refugee community(i.e. camp burns secondary to open-flame cooking) [16, 17].

Few studies have examined the physical trauma and injury burden sustained by refugee communities, particularly in the Eastern Mediterranean Region (EMR) [15, 18]. Global health has traditionally focused on communicable diseases with limited attention dedicated to injuries [19]. The main objective of this study is to quantify and describe the injury burden among a refugee community in the Northern Beqaa region of Lebanon and offer insights into injuries characteristics, extent, risk factors and outcomes. Understanding the frequency and severity of refugee injuries is vital to the planning of injury prevention programs, mitigation of injury impact on the refugee community and for reducing the strain on the Lebanese healthcare system.

## Methods

### Study Design and Tool adopted

The study was designed as a cross-sectional cluster-based population survey. The Surgeons Overseas Assessment of Surgical Need (SOSAS) tool version 3.0 (www.surgeonsoverseas.org) was used to collect data from 21 refugee camps across the Bekaa Valley in Lebanon. This survey is a validated, cluster-based, cross-sectional tool designed based on the Demographic and Health Surveys (DHS) guidelines and the WHO Guidelines for Conducting Community Surveys for Injuries and Violence to determine the burden of surgical conditions within a community [20-24]. Minor modifications were applied to contextualize to the tool and tailor it to the Syrian refugee population and expand it to collect specific data on physical injuries and burns (Appendix A). For the purpose of this study, a burn injury was captured independently and not considered under the umbrella of physical injuries.

### Data Collection

The survey was administered to the head of the refugee household and recorded all injuries sustained by family members over the last 12 months. A total 750 households were conveniently sampled by selecting the 4th residence along the most accessible major routes. If the tent was vacant or absent of adult, the next residence along the route was selected. Participants provided verbal informed consent prior to their participation.

Four bilingual research assistants familiar with the local geography, culture and Arabic language were recruited and trained to administer the SOSAS questionnaire. Training data collectors was carried out over a period of two weeks and included simulated practice sessions with role-play followed by a supervised data collection process at camp sites for the initial phases of the study. Data collection took place over three months between February - April 2019.

The study was approved by the American University of Beirut Institutional Review Board (SBS-2018-0561).

### Statistical Analysis

The Statistical Package for Social Sciences (SPSS) v. 24 (IBM Corp., Armonk NY, USA) was used for data management and analyses. A descriptive analysis was performed. Continuous data were reported as means with standard deviations, and comparisons made using the independent t-test. Categorical data were reported as counts with proportions and comparisons reported using the Chi-Square test, or the Fisher’s exact test, as appropriate. A two-sided P-value <0.05 was used to indicate statistical significance.

Missing data was encountered secondary to an incomplete response rate for all questions. Only collected data were analyzed, and no imputation techniques were used for any missing responses. The percentage of missing data for each variable is underlined in the footnote of its respective table.

## Results

### Demographics

A total of 750 heads of household were surveyed. The mean number of individuals per household/informal tented settlement (ITS) was 6.4. The mean age of individuals in each household was 20.4 years (Figure 1). Syrian governorates of origin were diverse with Aleppo (31.7%) and Raqqa (30.0%) being the most highly represented region, followed by Homs (12.2%), Idleb (11.3%), and Hama (7.5%). The average length of stay in Lebanon was 5 years (+/- 2.5). The majority of refugees were illiterate (59.0%) and nearly (72.2%) were unemployed.

**Figure 1:**
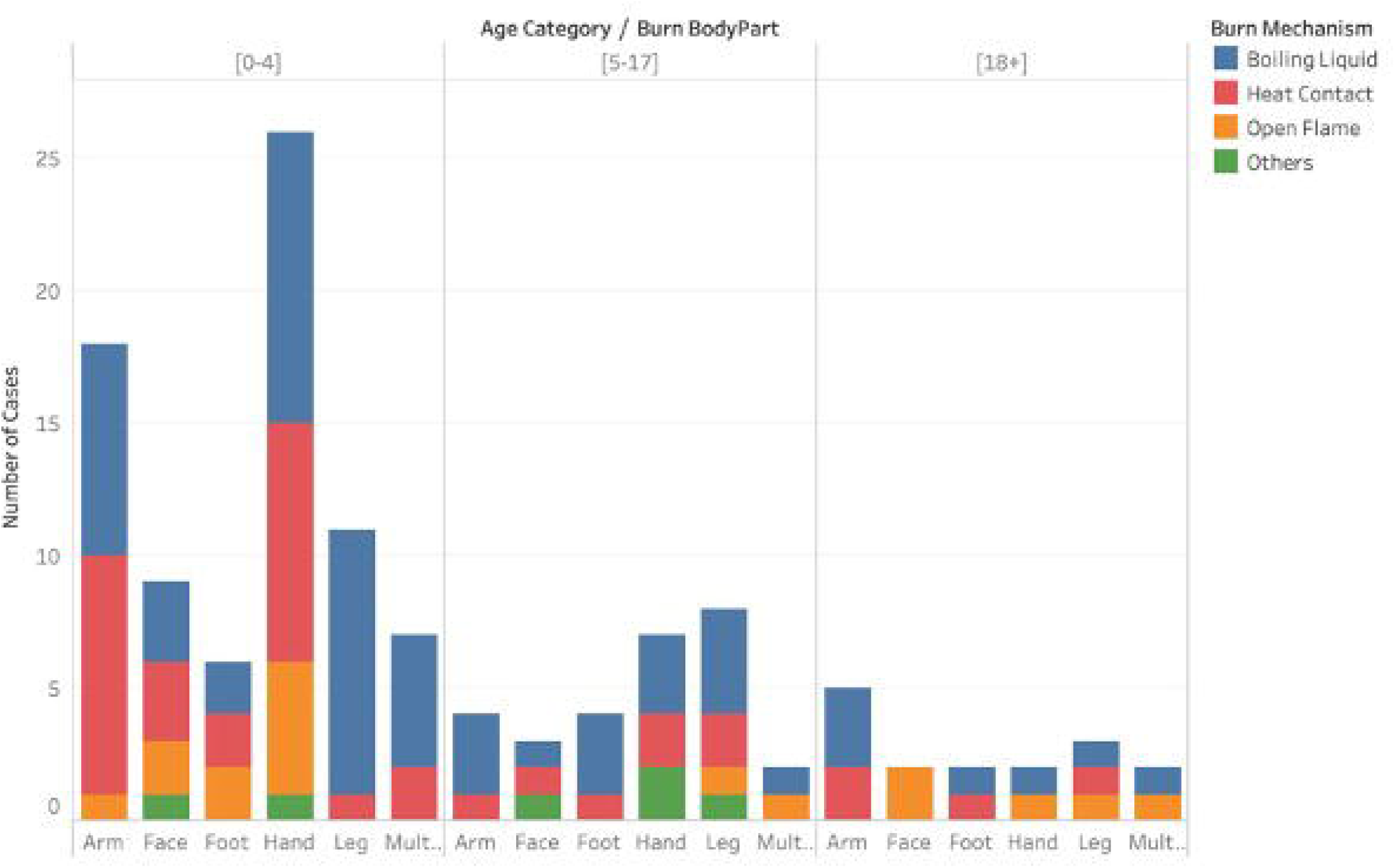
Distribution of injury across age, mechanisms and body parts

### Prevalence of Physical Injury

A total of 112 households (14.9%) reported sustaining an injury within the past year. Of these 18 (16.1%) reported being unable to seek health care services due to lack of financial means.

Injuries were sustained at multiple locations with the highest being inside the tent (29.9%) and on the road (28.6%) (Table 1). Twenty-two (19.6%) injuries were classified as occupational. Road traffic injuries (RTI) were sustained in 19 households (17%), most of which (73.7%) were due to a motorcycle crash. None of the injured refugees were adopting any safety measures while driving.

**Table 1.** Injury distribution on different locations

| Location | Number (%) |
| --- | --- |
| **On camp (Inside Tent)** | 23 (29.9) |
| **On camp (Outside Tent)** | 10 (13) |
| **Road** | 22 (28.6) |
| **Field** | 3 (3.9) |
| **Other** | 19 (24.7) |

The long-term effect of injuries varied, with 70 (62.5%) injuries reported as non-disabling, 39 (34.9%) injuries affected individuals’ work and daily living, and 3 (2.7%) injuries were reported as having a psychological impact (i.e. feeling ashamed) (Figure 2).

**Figure 2:**
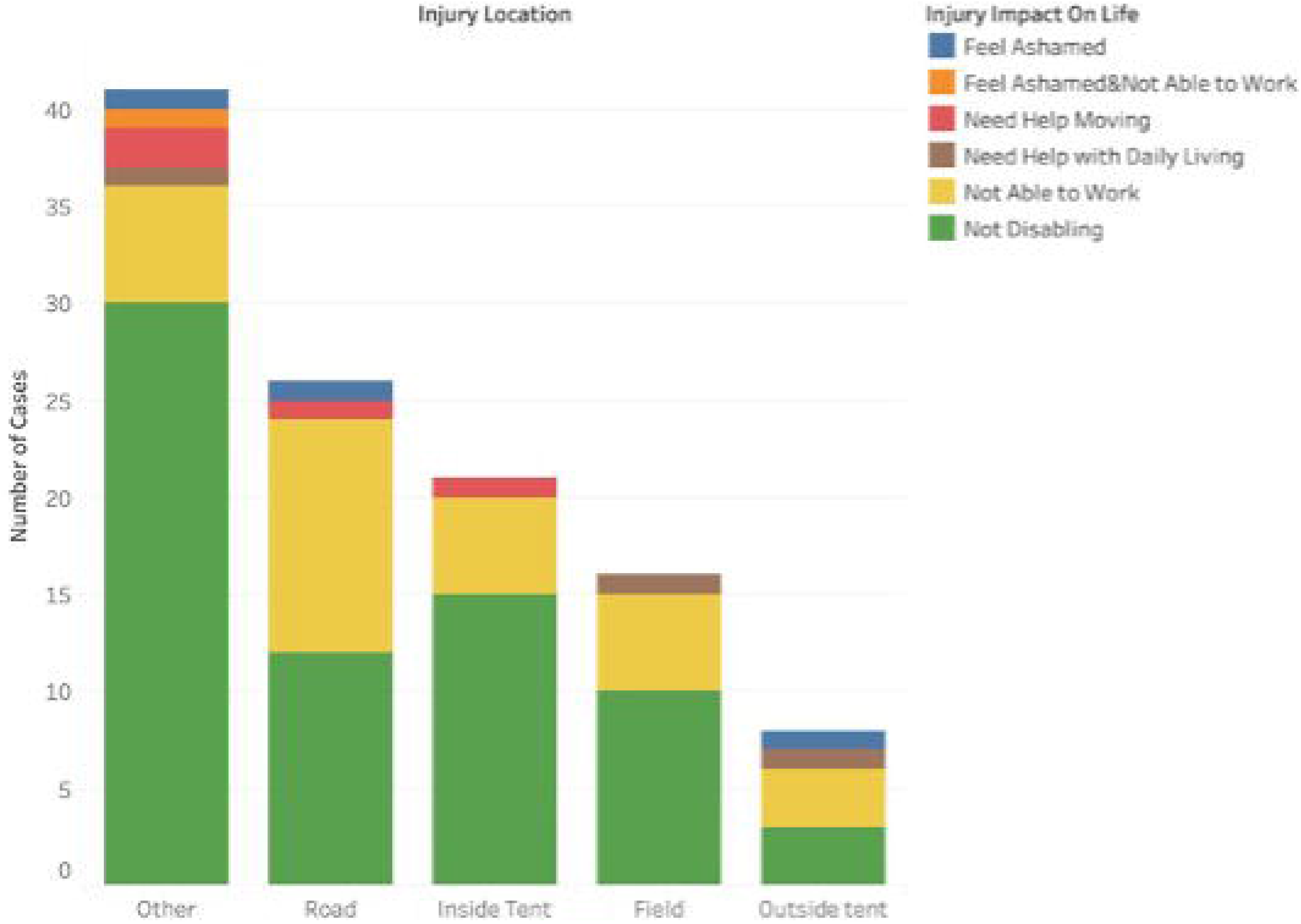
Distribution of injuries by type and impact on life

### Prevalence of Burns

A total of 136 households (18.1%) reported a burn to one of the household/tent members. Of those suffering from burns, 67 (53.6%) were male and 58 (46.4%) were females. The mean age of the injured individual was 8.2 ± 12.4 years and the majority (63.7%) of burns were incurred by children less than 5 years. Burns affected different body parts, with the highest being the hand/arm (50%), leg (19.4%) and face (11.3%) (Table 2).

**Table 2.** Burns distribution on body parts

| Body Part | Number (%) |
| --- | --- |
| **Hand** | 36 (29) |
| **Face** | 14 (11.3) |
| **Arm** | 26 (21) |
| **Leg** | 24 (19.4) |
| **Foot** | 13 (10.5) |
| **Multiple** | 11 (8.9) |

\*12 burns unknown

Half of the burns (49.6%) were caused by direct contact with a boiling liquid, 30.9% by contact with hot objects, and 13.8% by contact with an open flame. The mode and location of cooking varied amongst households While 78.4% of households use propane for cooking, 21.6% use open flame. Most cooking occurs inside tents (77%). The chi-squared test showed significance when examining Burn and No Burn among refugees’ households. There was no significant association between the mode or location of cooking and sustaining a burn injury (Table 3). Of the 750 households, 227 (30.3%) reported the inability of child lockout while cooking. Those tents had significantly higher reported burn cases (25.6% vs 14.9%, p =0.001). Households with an unemployed head member were more likely to sustain more burns (19.7% vs 13.3%, p =0.05) compared to employed households (Table 3).

**Table 3.** Incidence of burns in association with different tent-related variables

| Variable |  | Number (%) |  | *p* value |
| --- | --- | --- | --- | --- |
|  |  | Burn | No Burn |  |
| Tents |  | 136 | 614 |  |
| **Mode of Cooking** | Propane | 79 (18.2) | 356 (81.8) | 0.204 |
|  | Open Flame | 28 (23.3) | 92 (76.7) |  |
| **Location of Cooking** | Indoor | 78 (18.2) | 351 (81.8) | 0.187 |
|  | Outdoor | 30 (23.4) | 98 (76.6) |  |
| **Child Locked** | Yes | 78 (14.9) | 445 (85.1) | **0.001** |
|  | No | 58 (25.6) | 169 (74.4) |  |
| **Employed Household** | Yes | 24 (13.3) | 147 (86.7) | **0.05** |
|  | No | 112 (19.7) | 457 (80.3) |  |
| **Literate Household** | Yes | 36 (14.8) | 207 (85.2) | 0.102 |
|  | No | 100 (19.7) | 407 (80.3) |  |

\*195 tents have unknown mode of cooking
\*193 tents have unknown location of cooking

## Discussion

This study examined the characteristics, risk factors and outcomes of injuries sustained by the refugee community in Lebanon. Refugee status increases individuals’ risk of sustaining various injuries, particularly injuries associated with overcrowded living conditions and hazardous work environments. This study provides evidence on the prevalence of injuries and burns among refugees to inform tailored and data-driven injury prevention programs and strategies applicable to the context of refugees in an attempt to mitigate the injury burden on the refugee community and to reduce the demand for health and rehabilitation services on host healthcare systems [16, 25, 26].

This study aligns with existing literature and confirms the high prevalence of injuries among the refugee population in Lebanon [18, 27, 28]. A recent local study indicated that a high proportion of medical care services provided to adult Syrian refugees is related to injuries compared to local residents Lebanon [18, 27, 28]. Moreover, injuries were the most common reason for hospitalization among refugees, accounting for nearly 19.8% of hospitalizations compared to 14.9% among the host community [29]. This discrepancy in injury rates has been documented in various countries; a Canadian study reported an increased rate of motor vehicle injuries, poising, suffocation, and overall injury-related hospitalization and mortality among refugees [27]. A similar study conducted in Denmark revealed high rates of fatal injuries among refugees [30].

Household injuries were mostly reported in this study. The majority of refugee injuries occurred in tents (29.9%), further highlighting refugees’ suboptimal housing conditions, overcrowded households and adjacently installed tents with minimal safety standards. Diminished housing conditions and the absence of safety measures in camps are major contributors to the high prevalence of injuries among refugees [17]. Road Traffic Injury (RTI) (17%) represented another major contributor to the refugee injury burden. A similar estimate (19%) of RTI was reported among Afghan refugees in Pakistan [31]. The adoption of safety measures (e.g. wearing helmets, safety gears) were near absent among injured individuals.

Limited access to healthcare services hinders refugees’ ability to obtain timely care following an injury. The large number (16%) of refugees who reported an inability to afford injury-related treatment underscores refugees’ social and economic disadvantages and their impact on health. Refugees often suffer from a lack of knowledge on how to navigate the healthcare system in host countries, and how to benefit from available health services [32, 33], which ultimately adds yet another barrier healthcare access.

With a fragile, highly privatized, and under-resourced healthcare system in Lebanon, providing care to the local population is already a challenge that is exacerbated by the refugee crisis [34]. This under-resourced healthcare system, particularly in refugee areas, coupled with refugees’ increased need for healthcare services beyond that of the local population, exacerbates the economic burden of refugee communities on the host healthcare system [35, 36]. Refugees are forced to cover their own health expenses, and often resort to borrowing money to cover their out-of-pocket expenditure for injury treatment.

Findings from this study confirms the high prevalence of serious injuries leading to varying levels of physical impairment that affect refugees daily living activities. Many of the reported injuries result in severe prognoses, which may lead to permanent disabilities. This would limit the integration of refugees into workforce and accordingly limit their financial capabilities, further increasing refugees’ burden on host countries. A study in war-torn Baghdad found the rates of permanent disabilities following unintentional injuries were as high as 56% [28]. A similar trend was found in the United Kingdom among a population of refugees and migrants with 38% of head injuries causing persisting disability [37].

Similar to physical injury, burns were also a common health problem among refugees with a prevalence of over 18%. This rate is comparable to other refugee populations: 11% among Afghan refugees in Pakistan, 17% among Syrian refugees in Turkey, and 7% among Syrian refugees in Belgium [18, 31, 38]. Refugee parental education level (e.g. illiteracy), cultural practices (e.g. child supervision, cooking traditions), and housing conditions (e.g. overcrowded, unsafe heating techniques) are known to be risk factors increasing the risk of sustaining injuries among refugees [39, 40]. Camps are often used long beyond their temporary design intention leading to structural failures that often compromise safety. Results of this study show that unemployment and inability to keep children away from cooking areas are associated with higher prevalence of burns. It is notable that the number of burn injuries was higher in households adopting unsafe cooking practices using open flames instead of propane, however this association was not statistically significant. Similarly, burn case numbers were higher among households where the head of household claimed illiteracy, but again, this was not statistically significant.

Based on the study findings, a series of recommendations to help reduce and control injuries among refugees can be proposed. First, refugee camps should be designed with high safety standards focused on avoiding injuries and burns (e.g. larger lot for each tent to reduce family overcrowding, build camps away from major highways to reduce RTI). Second, a special focus should be given to the safe placement of heating and cooking appliances within camps. A tailored training on safe cooking practices should be considered.

Third, adequate occupational health and safety (OHS) training should be provided, with a focus on industrial and other high-risk work environments. Finally, refugees should be educated on how to access the local healthcare system and methods for obtaining financial support for health-related needs.

To our knowledge, this is the first study to quantify physical injuries and burns among Syrian refugees in Lebanon which has the highest Syrian refugee per capita density. The results of our study can be generalizable to other refugee populations in the MENA region, which share many cultural practices and living conditions. This study has several notable limitations. First, data is largely self-reported by household members. Recall bias must be considered as the collected information spans over a twelve-month period. Second, data underreporting is considered another possible limitation of this study. This, however, might be mitigated by social concerns and fear of stigma that may lead to underreporting of injury-related disabilities, particularly those affecting women and children.

## Conclusion

Refugees suffer from a high burden of injury and burns in Lebanon, with substantial human and economic repercussions on families and the host healthcare system. This study provides evidence to inform injury prevention programs affecting refugees. Education on safety guidelines and preventive measures should be introduced as standard protocol among the refugee community. Resources should be allocated for safe camp design, with a special focus on heat appliances and cooking methods. Further research is needed to understand the circumstances surrounding various types of injuries among refugees.

## Data Availability

Data will be available upon request.

